# Disentangling Socioeconomic Status and Race in Infant Outcomes: A Neural Network Analysis

**DOI:** 10.1101/2021.12.21.21268208

**Authors:** Kathryn Sarullo, Deanna M. Barch, Christopher D. Smyser, Cynthia Rogers, Barbara B. Warner, J. Philip Miller, Sarah K. England, Joan Luby, S. Joshua Swamidass

## Abstract

Race is commonly used as a proxy for multiple features including socioeconomic status. It is critical to dissociate these factors, identify mechanisms that impact infant outcomes, such as birthweight, and direct appropriate interventions and shape public policy. Demographic, socioeconomic, and clinical variables were used to model infant birthweight. Non-linear neural networks better model infant birthweight than linear models (R^2^ = 0.172 vs. R^2^ = 0.145, p-value=0.005). In contrast to linear models, non-linear models ranked income, neighborhood disadvantage, and experiences of discrimination higher in importance while modeling birthweight than race. Consistent with extant social science literature, findings suggest race is a linear proxy for non-linear factors. The ability to disentangle and identify the source of effects for socioeconomic status and other social factors that often correlate with race is critical for the ability to appropriately target interventions and public policies designed to improve infant outcomes as well as point out the disparities in these outcomes.

## Introduction

Socioeconomic status (SES) is currently the most robust predictor of child developmental and health outcomes (1–3). Experiences of deprivation and trauma known to be associated with low SES play a key role in the process by which adversity negatively impacts brain development and health behaviors (4–7). The drivers of racial disparities in health remain less well understood, in part due to the high collinearity between race and SES in many study samples, particularly those in the US, making it difficult to distinguish these effects from each other and often leading to race being determined as an essential risk factor for health in medicine. In addition, members of minority racial groups experience forms of discrimination and related obstacles that bring unique psychosocial stresses, as well as decreased access to necessary services and opportunities. These types of structural and social stressors have deleterious effects on health trajectories (8). Despite the complexity of these interrelationships, it has become clear from a variety of carefully conducted studies that accounting for SES attenuates the relationships of race to health outcomes (9). However, in previous work that used traditional statistical methods, race and low SES often interact because of the compounding of structural racism with economic disadvantage in the US, leading to associations between race and health even after adjustment for SES (10,11). More work is needed to elucidate and identify the factors that account for these residual relationships of race, particularly various forms of discrimination, trauma, and adversity.

Machine learning (ML) demonstrates superior modeling performance in a variety of domains, such as imaging, text analysis, genetics and more. Due to this, ML has begun to revolutionize the field of medicine (12,13). Linear regression (LR), or other more common statistical methods like Structural Equation Modeling (SEM), are often preferred by the medical community because of greater ease of interpretation. However, LR and SEM methods only work well when the underlying relationships among variables of interest are linear. Machine learning, specifically neural networks (NN), can be as interpretable, expose important non-linear relationships in the data, and reveal structures otherwise missed that may contain critical information. Additionally, NN allow the inclusion of multiple highly correlated variables without reducing performance and without loss of robustness. The goal of the current analysis was to build an interpretable neural network that models the effects of birthweight, one of the very first indicators of later health (14–19) and developmental outcomes with extremes of birthweight, such as small for gestational age (SGA; <10^th^ percentile at birth) and large for gestational age (LGA; >90^th^ percentile at birth), established as sensitive markers of cardiometabolic and neurodevelopmental risk into adulthood (14,15). Critically, data also suggest a relationship between birthweight within the normative spectrum to later childhood cognitive outcomes (16–18) and adult cognitive, educational, and earning achievements (19). Furthermore, this analysis will quantify the contribution of variables to the predictive power of models. The use of NN to accomplish these aims could have a high payoff by using the available data to understand the social determinants of infant outcomes.

In the current study, we sought to investigate the differential relationship of SES and race, as well as other forms of adversity to the mother, to fetal development during pregnancy in a study of the social determinants of health called “Early Life Adversity and Biological Embedding of Risk for Psychopathology” (eLABE) (20). Previous work from this group using SEM demonstrated the central relationship of Social Disadvantage, a latent factor that included income-to-needs, insurance status, education, area deprivation, and maternal nutrition, to birthweight. While SEM is valuable in its ability to determine relationships between variables, it remains limited by its requirement of linearity. It was unable to demonstrate dissociable relationships of race and SES to birth outcomes (20), due likely to high collinearity between SES and race in the sample as is common in many study samples worldwide. Drawing from prior work in social epidemiology and population health, we hypothesize that race often serves as a proxy for complex effects of social and economic disadvantage, and that such interrelationships may be non-linear, and thus often difficult to disentangle in studies using only linear statistical methods (21–25). Based on this finding and the central importance of the question, we investigate the utility of NN in disentangling the relationships of race and social adversity to infant outcomes versus a more standard linear regression approach.

## Results

The non-linear model accounted for more of birthweight variance than the linear model (R^2^ = 0.172 vs. R^2^ = 0.145, p-value=0.005) (Figure 1). P-values here were calculated using a comparison of correlations from dependent samples described in Statistical Analyses (26).

**Figure 1:**
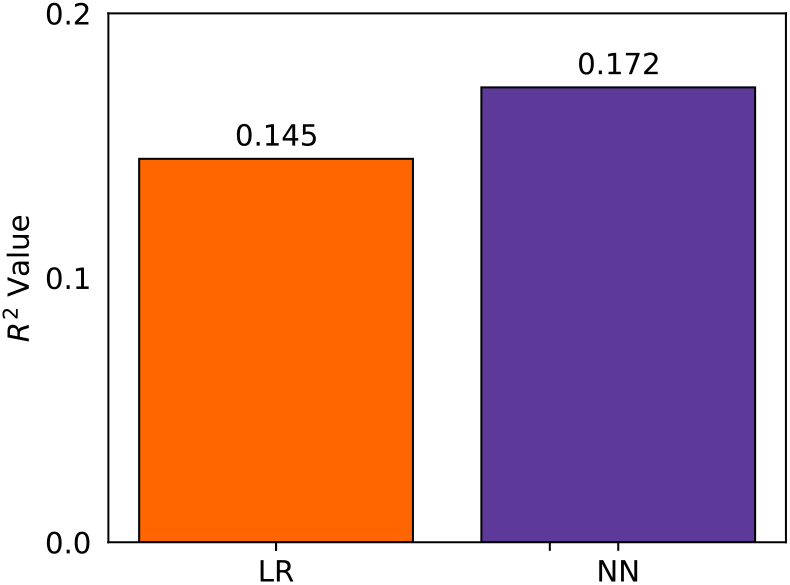
The non-linear model better fits the data (cross-validated R^2^ = 0. 172 vs. R^2^ = 0. 145, p-value=0.005 (26)). This improvement in performance is robust and repeatable across several cross-validation splits and training protocols. This improvement over the linear models – 2.7% absolute R^2^ increase and 18.6% relative R^2^ increase – indicates there are important non-linear relationships that the NN exploits.

### Feature Importance

We empirically quantified the contribution of each variable to model performance (Figure 2). The two most important variables for modeling birthweight, in both the linear and non-linear models, were maternal medical risk and maternal BMI (Figure 2A). In contrast to the linear model, the non-linear model was less reliant on the race variable in predicting birthweight (Figure 2A). The linear model ranked race as the next most important variable, followed by maternal income and neighborhood disadvantage. In contrast, the non-linear model ranked area deprivation, discrimination, and income as more important than race.

**Figure 2:**
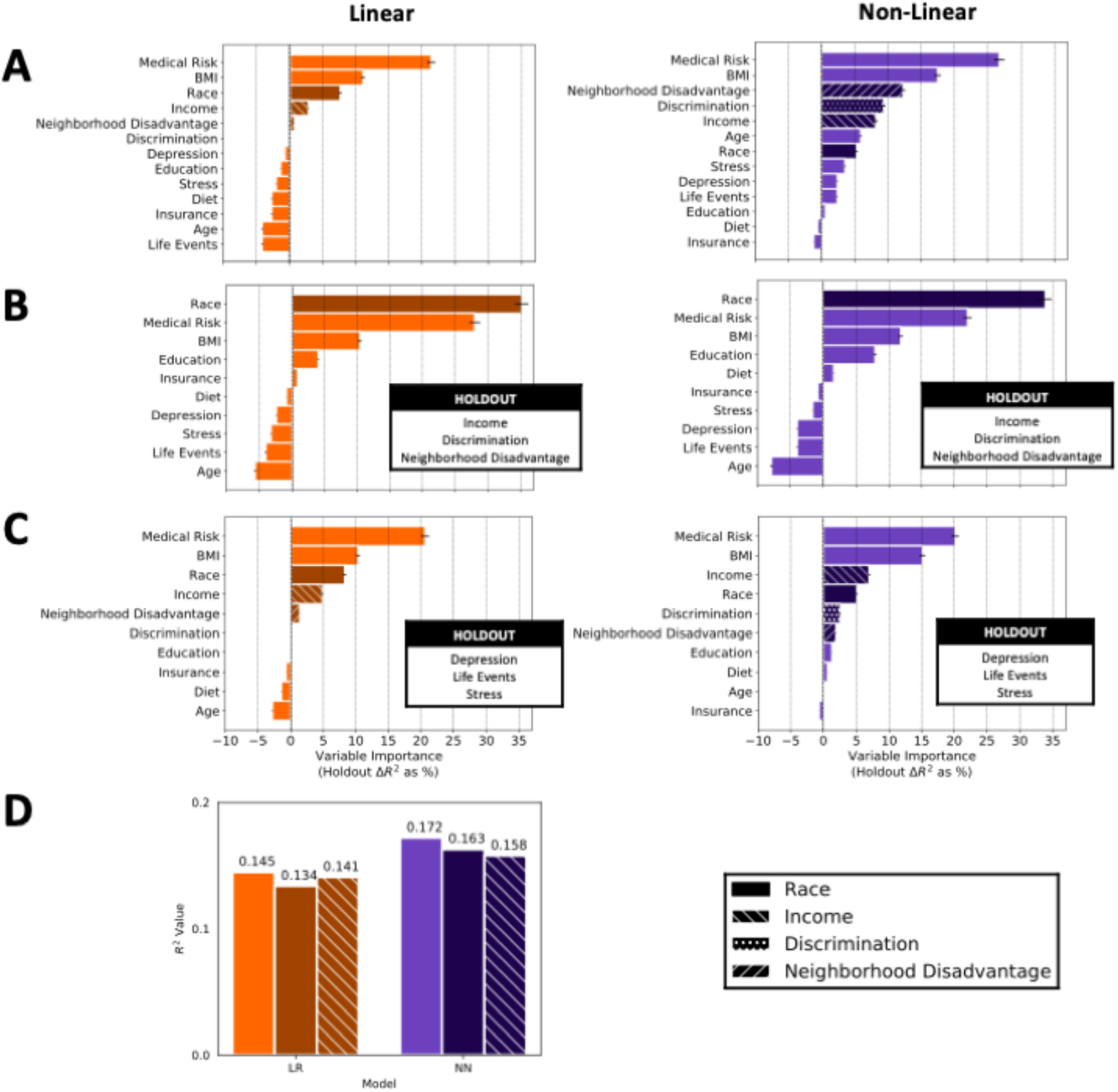
Race is less important in the non-linear model. Feature importance is robustly quantified by measuring the difference in R^2^ performance in models trained with and without the variable in question. (A) The non-linear model makes use of more variables than the linear model. Race is of reduced relative importance (3^rd^ vs. 7^th^ greatest effect) and is of reduced absolute performance (7.58% drop vs. 5.23% drop). Notably, income and discrimination variables are interrelated with race, and serve as a control, showing changes in the opposite direction as race in the non-linear model. (B) Both non-linear and linear models rely more heavily on race when income, discrimination and neighborhood disadvantage are held out. (C) In a negative control experiment, the impact of race did not decrease when stress, depression, and life events were held out. (D) The R^2^ performance degrades more with the removal of income (R^2^ = 0.172 vs. R^2^ = 0.158).

When holding out socioeconomic variables, the non-linear model becomes more dependent on the race variable (Figure 2B). Specifically, a holdout experiment was performed where household income, neighborhood disadvantage, and the discrimination survey were removed entirely (Figure 2B) from both models. Without those factors in the model, both the linear and non-linear model are more reliant on the race variable.

Holding out other variables did not increase the non-linear model’s dependence on race over income (Figure 2C). Specifically, as a negative control, we held out stress, depression, and life events (Figure 2C). We found that income is still ranked higher than race by the non-linear model, while in the linear model race continues to be higher ranked than income.

The linear model relies more on race while the non-linear model relies more on income to predict birthweight. Additional information is derived from the single-variable holdout R^2^ performance of each model (Figure 2D). The linear model has a larger decrease in R^2^ when race is removed, perhaps because it is linearly correlated to the infant outcome and it relies on that information for predictive performance. NN has a larger decrease in R^2^ when income is removed, perhaps because there is a non-linear correlation between income and infant outcomes.

### Non-linear Responses and Interactions

The difference in performance and variable importance between NN and LR appears to be due to several subtle non-linear relationships. To better understand why the non-linear model outperformed the linear model, we first examined non-linear relationship of the predictors to the outcome. There are very subtle non-linear relationships of the various predictors to birthweight within the model (Figure S1A). The most non-linear univariate response was to BMI (RMSD (g) = 17.63) and depression (RMSD (g) = 14.94). The non-linear model also captured several non-linear interactions (Figure S2A, Table ST1). The largest non-linear interaction was between depression (RMSD (g) = 32.75) and life events (RMSD (g) = 16.44) (Figure S2B, Figure S2C). Therefore, the non-linearities modeled are individually subtle; yet, collectively, they dramatically impacted the empirical importance of input variables.

## Discussion

The goal of this work was to determine if non-linear neural networks could better dissociate the relationships of race versus a range of psychosocial and biological factors known to impact health, specifically birthweight, a key outcome known to be broadly predictive of health trajectories (14–19). We found that the non-linear model outperformed the linear model in terms of the amount of variance accounted for in infant birthweight. This suggests that there are non-linear relationships within these data that the NN can exploit while the linear model cannot. Though the non-linearity was subtle, meaning a relatively small RMSD in grams, the impact on variable importance was robust and large. This result was reproducible and implies that there are non-linear relationships not being captured by a linear model, however small they may be. It is common to employ linear models after ensuring there are no clear non-linear relationships between variables and the outcome variables. Our results show, however, that even subtle non-linear effects can have a large impact on assessments of variable importance and in being able to disentangle variables relevant to the social determinants of infant health.

These findings have important implications for the understanding of which elements of the global experience of psychosocial adversity are driving risk for poor health outcomes. Refuting prior work that has made assumptions about the biological role of race, these findings demonstrate that race is acting as linear proxy for a variety of non-linearly correlated adverse experiences, including economic disadvantage and discrimination (21–25). Here we show these relationships and associations to infant birthweight, one of the very first indicators of later health (14–19) and developmental outcomes associated with brain development (14,15), and later childhood cognition function (16–18) and adult cognitive, educational, and earning achievements (19). As such, there may be significant value in testing non-linear variable importance in clinical studies of other components of the social determinants of health. Perhaps with non-linear modeling, the mechanisms of disparate outcomes correlated with race can be better understood.

These findings about the importance of income and discrimination have very powerful public health implications for the design of prevention programs that should be targeting discrimination and other forms of social adversity related to low SES. They also elucidate the importance of these factors during pregnancy and their effects on birth outcomes. These data highlight the importance of pregnancy as a key window of time for future health preventive interventions targeting the child.

In terms of limitations, this dataset for this study is small compared to many studies that use neural networks. Due to this limitation, we used a rather simplistic neural network with only one hidden layer with a width of 2 as opposed to a deep and wide neural network. However, the same size for the current study is relatively large for an infant outcome study. Further, it will be important to replicate these findings in additional data sets, such as those being collected as part of the Environmental Child Health Outcomes (ECHO) study or the upcoming Health Brain Cognitive Development Study (HBCD).

These results disentangle the relationships between race and SES to better inform interventions and public policy. It uses data about experiences of adversity and advantage during pregnancy and other factors to explore which variables were most important to one key infant outcome, birthweight.

Non-linear models better modeled infant birthweight due to non-linear responses and interactions within the data. In contrast to linear models, non-linear models ranked income, area deprivation index (ADI), and experiences of discrimination higher in importance than race. Even though the non-linearities modeled are individually subtle, the relative importance of variables can be importantly and robustly different.

A comparison of the linear and non-linear models is consistent with race acting as a linear proxy of a non-linear combination of other variables in this dataset. By modeling the non-linear relationships directly, non-linear models better disentangle the relationship between race and SES in this study of maternal adversity and birthweight. This will assist in understanding the disparities among the data and their origins. As such, these findings significantly extend our understanding of how to disentangle the complex relationships between race and SES in understanding maternal adversity and income outcomes by identifying non-linear relationships that reveal the importance of socioeconomic variables over race in predicting infant birthweight, one of the early outcomes of maternal adversity.

### Future Work

In future work, similar non-linear models and similar feature importance techniques can be used to examine relationships to other infant outcomes, such as brain structure and function at birth, potentially including birthweight and gestational age as input variables. In addition, future studies that employ experimental designs can explore disentangling groups of variables using causality. This could be very helpful in determining mediation or causality between those highly correlated variables and the output. More generally, it can be used to determine causality between any variable or set of variables a user wants to investigate with appropriate study designs.

In terms of limitations, this dataset for this study is small compared to many studies that use neural networks. However, the same size for the current study is relatively large for an infant outcome study. Simplistic techniques have been implemented to fill in missing data. To increase the size of the dataset, the existing data could be used to create synthetic patient data. More complex methods could be implemented, such as using available data to predict the missing variable. The size of the dataset could also be increased by using the existing data could be used to create synthetic patient data. There are tools that exist that create similar, yet different, patient data that resembles the existing data, hence increasing the size of your dataset.

In summary, this work begins to disentangle the relationships between race and SES to better inform interventions and public policy. It uses data about experiences of adversity and advantage during pregnancy and other factors to explore which variables were most important to one important infant outcome, birthweight. Non-linear models were able to better model infant birthweight due to non-linear responses and interactions within the data. In contrast to linear models, non-linear models ranked income, area deprivation index (ADI), and experiences of discrimination higher in importance than race. The non-linear model suggest that race is a proxy of a non-linear combination of other variables in this high-risk urban US study sample and that non-linear components are needed over linear to better model differential impacts on infant outcomes. As such, these findings significantly extend our understanding of how to disentangle the complex relationships between race and SES in understanding maternal adversity and income outcomes.

## Materials and Methods

The Early Life Adversity Biological Embedding and Risk for Developmental Precursors of Mental Disorders (eLABE) is a multi-wave, multi-method NIMH-funded study designed to investigate the mechanisms by which prenatal and early life adversity impact infant neurodevelopment. All study procedures were previously approved by the Washington University School of Medicine Institutional Review Board (WUSM IRB). Pregnant women who were participants in a large-scale study of preterm birth within the Prematurity Research Center at Washington University in St. Louis with negative drug screens (other than cannabis) and without known pregnancy complications or known fetal congenital problems, were invited for eLABE participation. The study recruited N=395 women during pregnancy (N=268 eligible subjects declined participation) and their N=399 singleton offspring (N=4 mothers had 2 singleton births during the recruitment period). Out of those originally invited and interested in participation, N=26, were deemed ineligible (N=13 screened out prior to consent and N=13 consented subjects were deemed ineligible due to later discovery of substance abuse or the finding of a congenital anomalies). Women facing social disadvantage were over-sampled by increased recruitment from a clinic serving low-income women. The sample was also enriched for preterm infants with N=51 born preterm (<37 weeks gestation). After removing participants that were missing entire surveys, this left 351 infants included in the current analysis (Figure 3).

**Figure 3:**
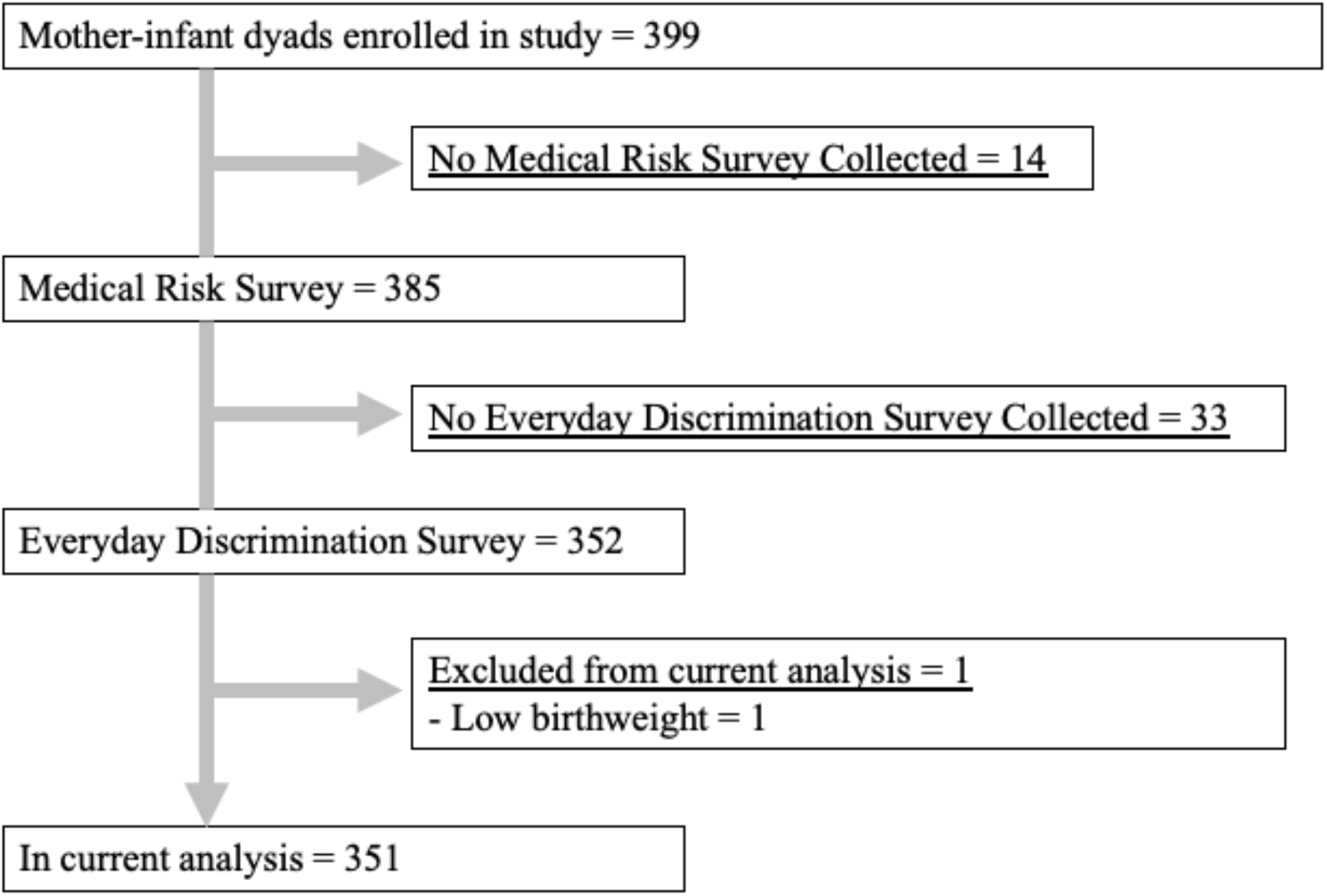
Participant flow from study enrollment to inclusion in current analysis. This flow diagram shows the number of initial participants and reasons for exclusion from this analysis.

### Data

As described, maternal depression, experiences of stress, as well as demographic information including insurance, education, address, and household composition were obtained from participants at each trimester during pregnancy. Maternal dietary and medical history were obtained from self-reported surveys and medical records while birthweight was obtained from delivery records. Mothers and their newborns were invited for an assessment shortly after birth which included neonatal MRI during which mothers completed a comprehensive measure of life stress and trauma (current and past) and discrimination (Table 1).

**Table 1:**
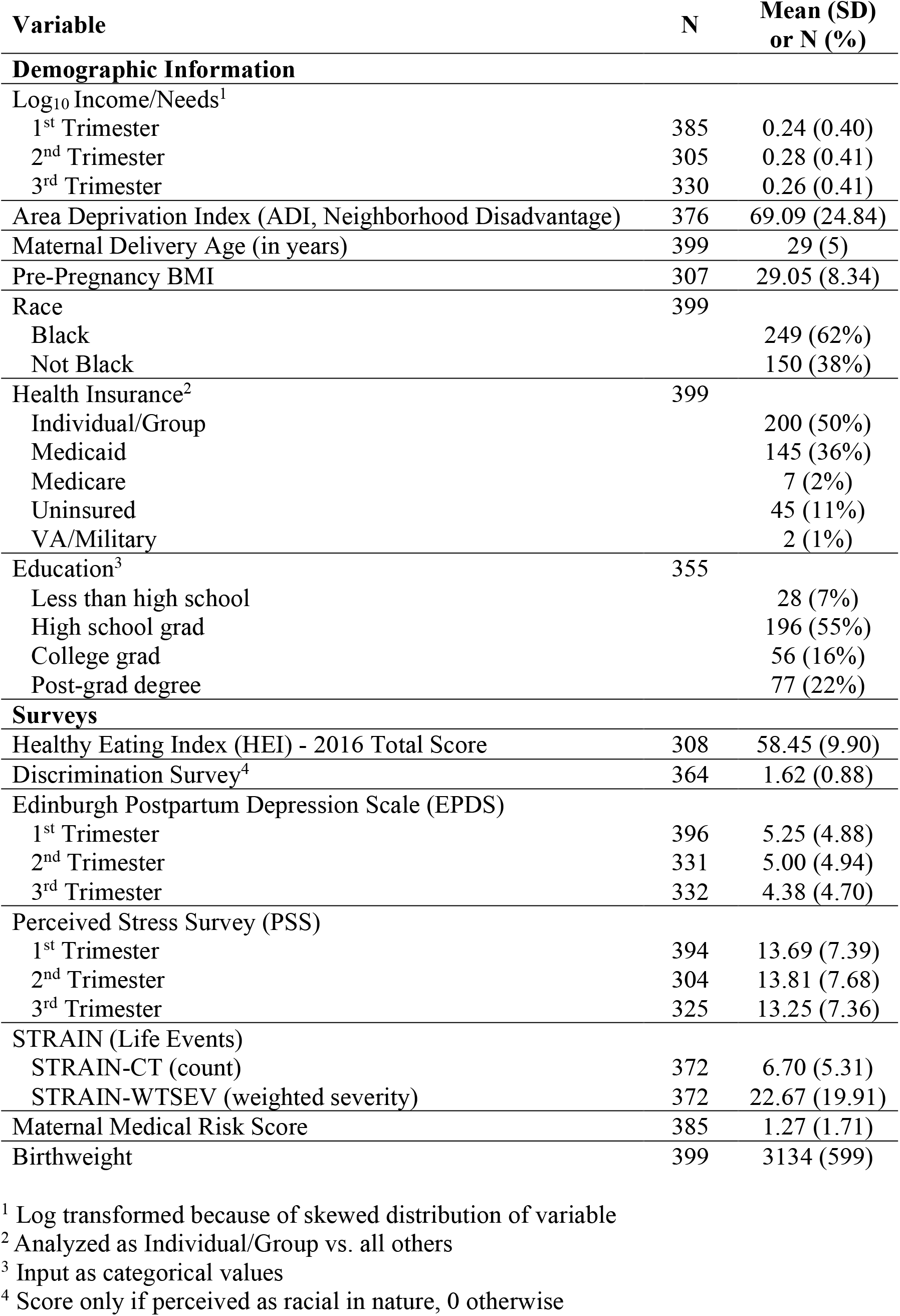
The aim is to model birthweight using these 20 variables.

| Variable | N | Mean (SD)<br>or N (%) |
| --- | --- | --- |
| **Demographic Information** |  |  |
| Log<sub>10</sub> Income/Needs<sup>1</sup> |  |  |
| 1<sup>st</sup> Trimester | 385 | 0.24 (0.40) |
| 2<sup>nd</sup> Trimester | 305 | 0.28 (0.41) |
| 3<sup>rd</sup> Trimester | 330 | 0.26 (0.41) |
| Area Deprivation Index (ADI, Neighborhood Disadvantage) | 376 | 69.09 (24.84) |
| Maternal Delivery Age (in years) | 399 | 29 (5) |
| Pre-Pregnancy BMI | 307 | 29.05 (8.34) |
| Race | 399 |  |
| Black |  | 249 (62%) |
| Not Black |  | 150 (38%) |
| Health Insurance<sup>2</sup> | 399 |  |
| Individual/Group |  | 200 (50%) |
| Medicaid |  | 145 (36%) |
| Medicare |  | 7 (2%) |
| Uninsured |  | 45 (11%) |
| VA/Military |  | 2 (1%) |
| Education<sup>3</sup> | 355 |  |
| Less than high school |  | 28 (7%) |
| High school grad |  | 196 (55%) |
| College grad |  | 56 (16%) |
| Post-grad degree |  | 77 (22%) |
| **Surveys** |  |  |
| Healthy Eating Index (HEI) - 2016 Total Score | 308 | 58.45 (9.90) |
| Discrimination Survey<sup>4</sup> | 364 | 1.62 (0.88) |
| Edinburgh Postpartum Depression Scale (EPDS) |  |  |
| 1<sup>st</sup> Trimester | 396 | 5.25 (4.88) |
| 2<sup>nd</sup> Trimester | 331 | 5.00 (4.94) |
| 3<sup>rd</sup> Trimester | 332 | 4.38 (4.70) |
| Perceived Stress Survey (PSS) |  |  |
| 1<sup>st</sup> Trimester | 394 | 13.69 (7.39) |
| 2<sup>nd</sup> Trimester | 304 | 13.81 (7.68) |
| 3<sup>rd</sup> Trimester | 325 | 13.25 (7.36) |
| STRAIN (Life Events) |  |  |
| STRAIN-CT (count) | 372 | 6.70 (5.31) |
| STRAIN-WTSEV (weighted severity) | 372 | 22.67 (19.91) |
| Maternal Medical Risk Score | 385 | 1.27 (1.71) |
| Birthweight | 399 | 3134 (599) |

<sup>1</sup> Log transformed because of skewed distribution of variable
<sup>2</sup> Analyzed as Individual/Group vs. all others
<sup>3</sup> Input as categorical values
<sup>4</sup> Score only if perceived as racial in nature, 0 otherwise

*<u>Income to Needs ratio</u>* (I/N) was measured at each trimester. The I/N ratio utilizes self-reported family income and household size compared to federal poverty thresholds, with a ratio of 1.0 being at the poverty line. *<u>Insurance status</u>* was collected at the time of enrollment through a medical record review and was verified in the third trimester or at delivery; *<u>Mother’s highest level of education</u>* was self-reported at the time of enrollment. *<u>Area Deprivation Index</u>* (ADI) or “neighborhood disadvantage” as we refer to it, is a geo-tracking measure used to rank neighborhoods by socioeconomic disadvantage compared to the national average based on census block data, including factors for the domains of income, education, employment, and housing quality (27,28). ADI is represented as a national percentile with a higher value indicating greater disadvantage. *<u>Maternal nutrition</u>* was assessed using the Healthy Eating Index (HEI) during the third trimester or at delivery. This is a validated dietary assessment tool available through the National Institutes of Health used to measure diet quality based on U.S Dietary Guidelines for Americans (29,30). Dietary information for HEI calculation was obtained using the Diet History Questionnaire (DHQII) (31,32). In each trimester, mothers completed the Edinburgh *<u>Postnatal Depression</u>* Scale (EPDS) (33) and *<u>Perceived Stress</u>* scale (PSS) (34). STRAIN (35), a comprehensive measure of *<u>lifetime stressful and traumatic life events</u>*, was collected at the time of neonatal scan (N=255) or at a follow-up exam (N=108); no differences in STRAIN scores based on time of administration were found. Experiences of *<u>discrimination</u>* based on race were assessed using the Everyday Discrimination Scale (36) measured at neonatal scan. To control for maternal medical risks that might be confounded with social or psychological disadvantage, we assessed *<u>maternal age at delivery</u>* and *<u>pre-pregnancy body mass index</u>* from first prenatal visit based on self-reported pre-pregnancy weight and height. In addition, a *<u>Maternal Medical Risk Score</u>* (MMR), containing pre-existing and pregnancy-related medical conditions, was computed using a validated measure of maternal medical comorbidities extracted from the medical record that accounts for 22 medical conditions weighted by severity (37). *<u>Birthweight</u>* was collected from the medical record at the time of delivery.

### Models

Linear regression (LR) is a widely used method that models data by fitting a linear equation, ŷ = mX + b.X is a matrix where every row represents specific patient information. The inputs are linearly combined to output the predictions, ŷ.

LR modeled birthweight as a linear combination of several clinical variables (Figure 4A). LR is a baseline for performance model since it is commonly used to model outputs in medicine. However, LR will only find linear relationships in the data. Likewise, LR will not fully control for non-linear confounding variables.

**Figure 4:**
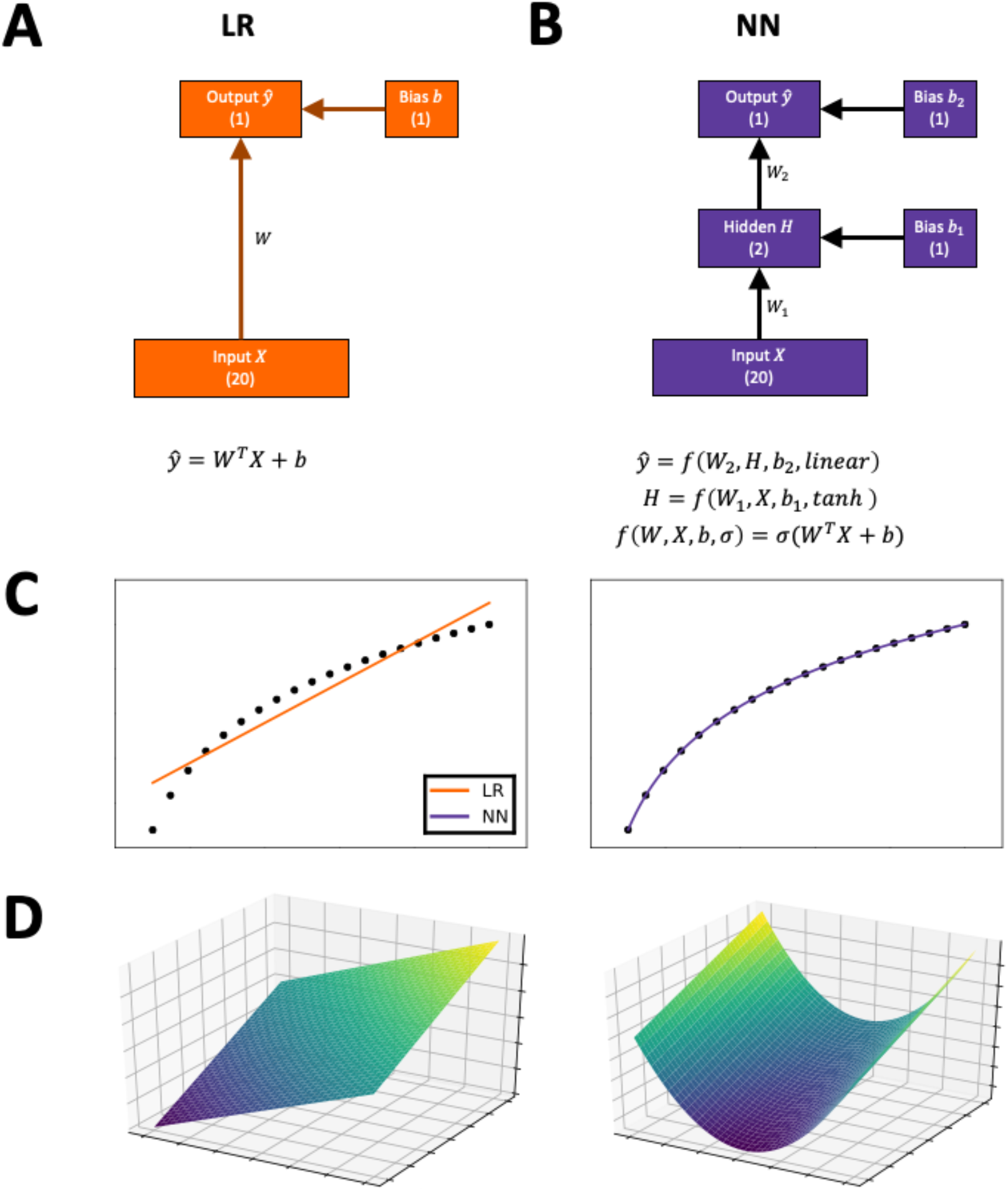
A depiction of the linear and non-linear models used in this study. Models are depicted in diagrams that show how the modeled birthweight is computed from input variables, with vectors depicted as boxes and arrows indicating flow of information. (A) The linear regression (LR) computes the output as a linear transform of the input vector. (B) The neural network (NN) transforms the input vector into a hidden layer of variables, and this layer is transformed into the output. In practice, models operate on normalized data, so all inputs and outputs are Z-normalized. (C) The NN can represent non-linear relationships, such as non-linear responses, better than LR can. (D) The NN can represent non-linear relationships, such as non-linear interactions, better than LR can.

Neural networks (NN) in machine learning make use of non-linear equations with more parameters. In the simplest form, each layer of a network is computed as, f(X) = σ(W^T^X + b), The data matrix X is linearly transformed by W and b, and then each element of the matrix is input to a non-linear function σ. Multiple layers, each with a different set of parameters, are cascaded to compute the final output. In this study, we use the tanh function as our non-linear function, and a single hidden layer (Figure 4B). Several other architectures were tested, but this one exhibited optimal performance (Figure S3).

NN are particularly useful and can perform equivalent to, sometimes exceeding, human ability to analyze and annotate imaging data (38). In contrast with LR, NN use non-linear functions to model non-linear relationships. We aim to test if NN will be more effective at controlling for non-linear confounders in this dataset.

NN are more complex than the LR, requiring more weights to be trained, but also have the possibility to outperform LR. For example, NN can model non-linear relationships between individual inputs and the target output (Figure 4C). Likewise, NN can model non-linear interactions between multiple inputs (Figure 4D). A NN can allow for improved correlation performance relative to LR if there are any such non-linear relationships.

### Statistical Analyses

We used the TensorFlow (39) Adam optimizer with 10-fold cross validation to train both LR and NN models. The TensorFlow error function mirrors ordinary least squares estimation, which is commonly used to train LR. All performance was computed using 10-fold cross-validated predictions. One tenth of rows were held out as validation set and the remaining observations are used as the training set. Ten models were trained with different hold-out sets, such that each observation was in the validation set once. Within each fold, each model was trained 20 times with a random restart, and the best performing model was automatically selected. This restart procedure ensured the results were robust. In cases where a specific patient datum was missing, the missing variable was filled in with the average value across all other patients for that variable.

Models were first compared by their respective cross-validated R^2^ performance, which describes the amount of variance accounted for while modeling birthweight. To calculate the p-values for significant differences between the R^2^ values, a test of correlation differences from dependent samples was employed (26).

For each model, we quantified which variables were important for predictive power. Individual variables or sets of variables were held out from the training data. The cross-validated performance on the reduced dataset was computed using the same protocol. The decrease in R^2^ quantifies the importance of the held-out variables. The larger the decrease, the more independent information that variable is contributing to R^2^ performance.

This procedure for measuring variable importance is related to a widely used approach called Shapley additive explanations or SHAP (40). SHAP is used to determine the importance of variables in machine learning algorithms by approximating SHAP values. These values explain how much an input variable impacts the output of model. However, when using non-linear models in SHAP, the results were not stable, producing very different results each run (Figure S4). In contrast, our approach measured the impact of variables on the global performance, not the model output, and yielded stable results across multiple runs.

To quantify non-linear responses and interactions, we used a clamping test. First, to measure univariate non-linearity, we considering a range of fixed values for the test variable. In turn, the input matrix was transformed by clamping the test variables at the given fixed value, and the average output of a trained model on the clamped dataset was computed. A non-linear relationship between the average output and the fixed value indicates non-linearity. We quantified non-linearity as the root-mean-squared-deviation (RMSD) of the best fit line of these measurements. A line has no non-linearity, so a larger RMSD indicates more non-linearity, which was used to rank and quantify the degree of non-linearity in individual variables.

Second, an analogous, bivariate non-linearity is computed in a similar way. Two variables are clamped at a range of values, covering a grid in 2D space. The input matrix was transformed to clamp the data of both variables at the given values, and the average model output is computed. quantified non-linearity as the RMSD of the best fit plane of these measurements. A flat plan has no non-linearity, so increased RMSD indicates more non-linearity in the interactions between two variables, and we used RMSD to rank pairs of input variables with the most non-linear response.

## Data Availability

All data produced in the present study are contained in the manuscript.

## Author contributions

Conceptualization: KS, SJS

Methodology: KS

Investigation: KS

Visualization: KS

Supervision: SJS, JL

Writing—original draft: KS, JL, DMB, SJS

Writing—review & editing: KS, JL, DMB, SJS, CDS, CR, BBW, JPM, SKE

## Data and materials availability

The data will be shared through NIMH with standard data sharing policy in effect

## Supplementary Materials

Please see supplementary file.

