## Supplemental_Data for "Disentangling Socioeconomic Status and Race in Infant Outcomes: A Neural Network Analysis"

**This PDF file includes:**

Figs. S1 to S4
Table S1

**A**

| Variable | RMSD (g) | R |
|---|---|---|
| BMI | 17.63 | 0.986 |
| Depression | 14.94 | -0.997 |
| Income | 13.24 | 0.996 |
| Discrimination | 11.42 | 0.980 |
| Medical Risk | 11.17 | -0.998 |
| Neighborhood Disadvantage | 7.86 | -0.995 |
| Age | 7.31 | -0.995 |
| Stress | 2.75 | -0.993 |
| Life Events | 2.56 | 0.997 |
| Diet | 1.78 | 0.999 |

**B**

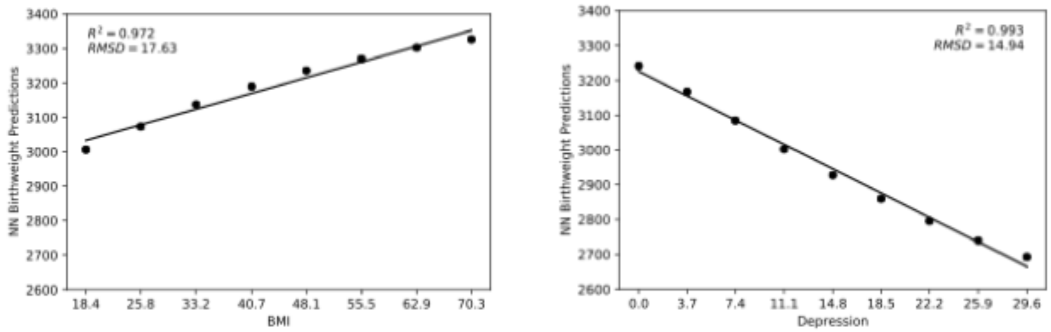


**Fig. S1. In contrast with the linear model, the non-linear model relies more on income and discrimination than it does on race.**

(A) The non-linear response was quantified for each variable. (B) Clamping the dataset's BMI at specified values shows a subtle non-linear response ($RMSD = 17.63$ vs. $RMSD = 0$ for a linear response), which the non-linear model may be relying upon. Similarly, clamping the depression survey total score shows a subtle non-linear response as well ($RMSD = 14.94$ vs. $RMSD = 0$ for a linear response). For comparison, the linear model does not model this non-linearity.

**A**

| Variable 1 | Variable 2 | RMSD (g) |
| --- | --- | --- |
| Depression | Life Events | 32.7501 |
| Depression | Discrimination | 16.4434 |
| Depression | Income | 16.2997 |
| Income | Life Events | 16.1404 |
| Depression | Stress | 14.5677 |
| Income | BMI | 13.9249 |
| BMI | Discrimination | 13.0348 |
| Depression | Medical Risk | 12.9667 |
| Income | Discrimination | 12.6559 |
| BMI | Life Events | 12.6086 |

**B**

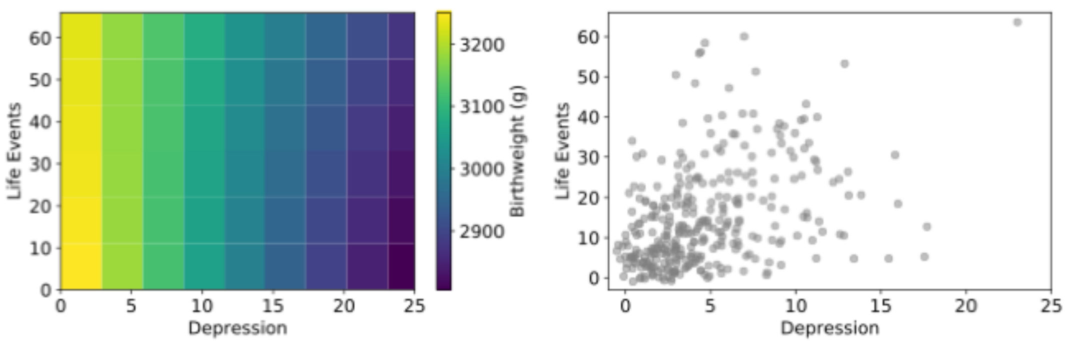


**C**

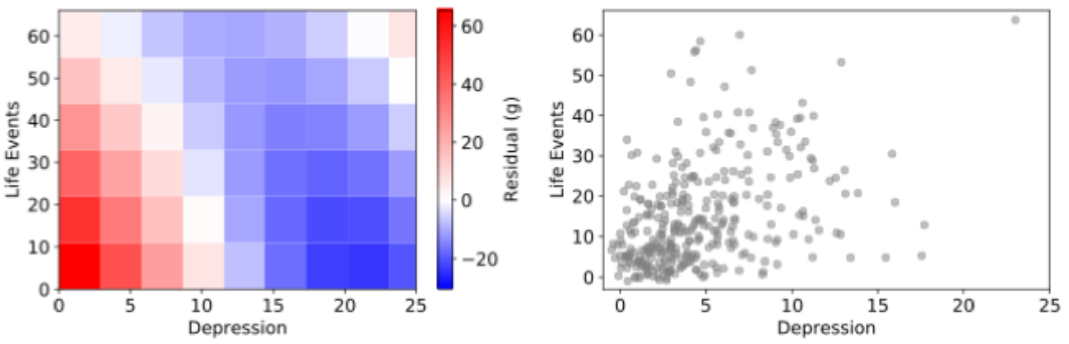


**Fig. S2. The largest non-linear interaction was between depression and life events.**
The clamping test was performed on two variables to calculate the interactions between combinations of variables. (A) The non-linear interactions were quantified between each combination of variables (Table ST1). (B) The largest non-linear interaction was between depression and life events. A heatmap shows the non-linear surface of their interaction. There is a subtle non-linear interaction here. The scatter plot of the actual datapoints help show where the important regions lie. (C) It becomes clearer to see the non-linear interaction by looking at the residuals between the non-linear surface and the linear plane are plotted in a heatmap. This is telling us that the combination of having a low depression score and low life events lead the non-linear model to predict a higher birthweight than the linear model. Another interesting example is the combination of having a high depression score and low life events leads to the non-linear model predicting a very low birthweight. The scatter plot of the actual datapoints help show where the important regions lie.

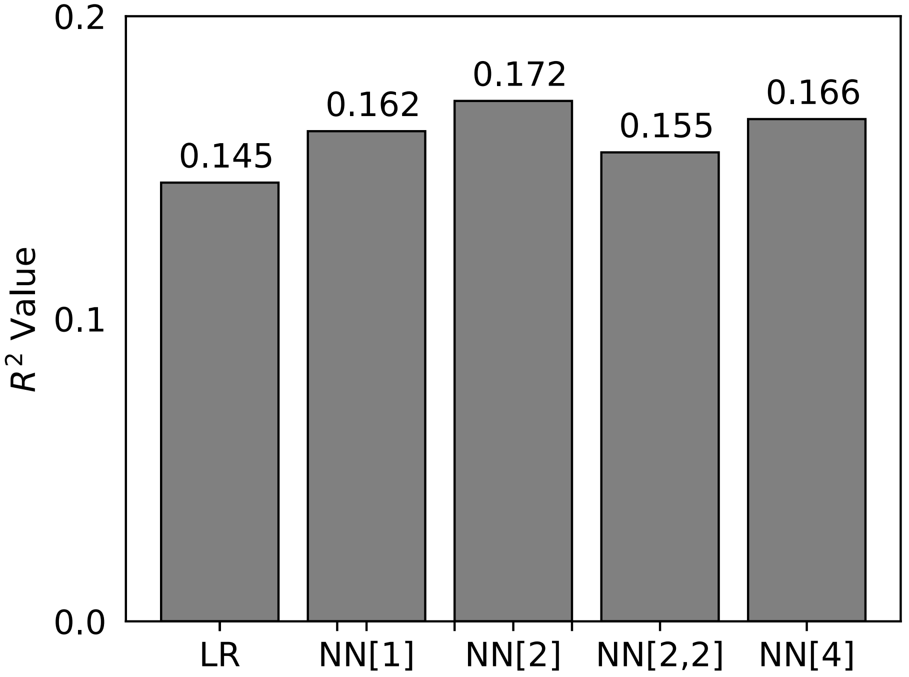


**Fig. S3. Performance of each model in terms of $R^2$.**

This shows the performance of the neural network with different architectures. We tried different widths for the hidden layer(s) of 1, 2 and 4 as well as adding a varying number of hidden layers, 1 or 2.

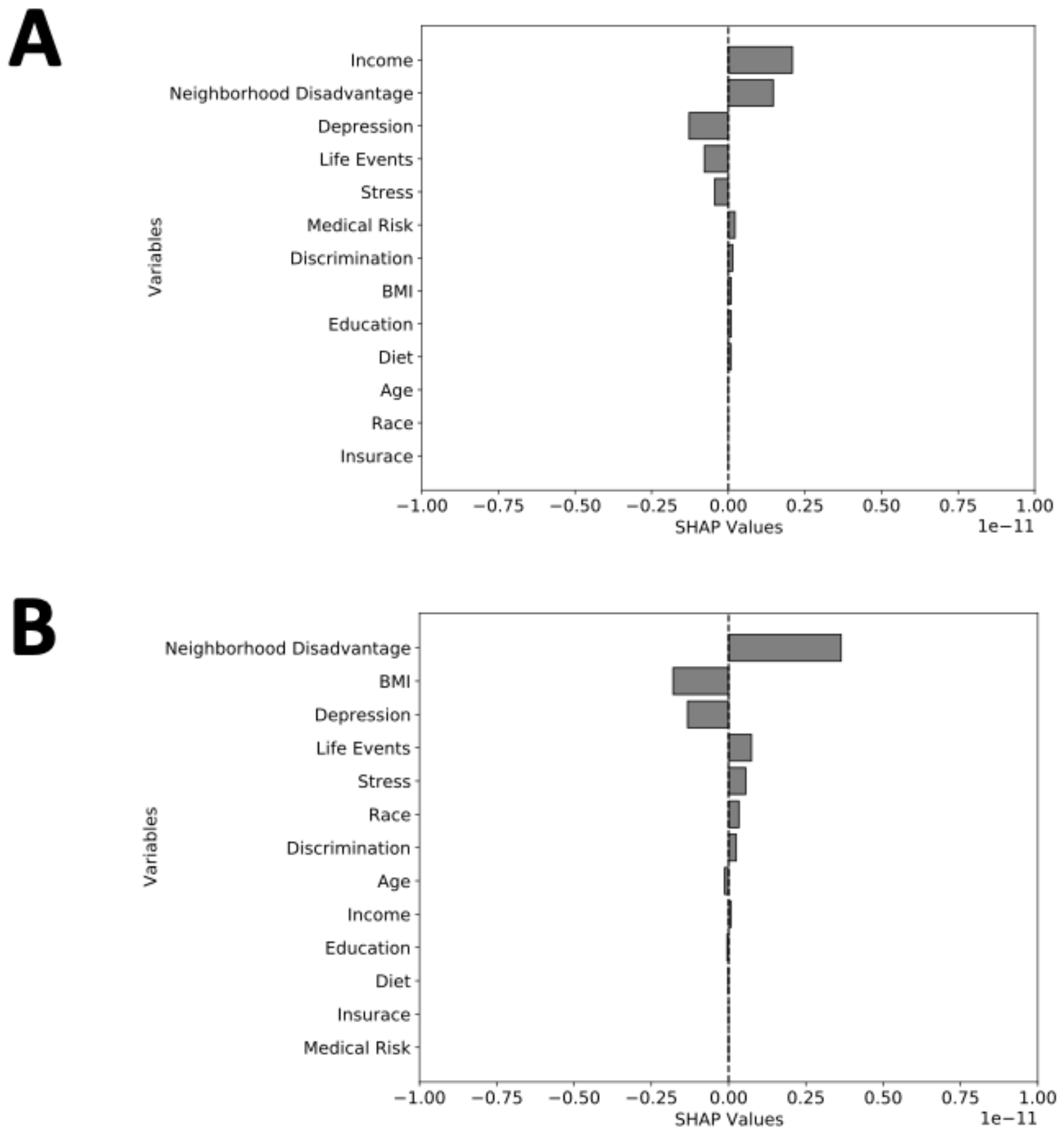


**Fig. S4. SHAP's DeepExplainer was not consistent from run to run using Keras with same architecture as NN.**

The SHAP values from run to run varied and were not robust and stable. (A) First run SHAP values. (B) Second run SHAP values.

| | | |
|---|---|---|
| Depression | Life Events | 32.7501 |
| Discrimination | Depression | 16.4434 |
| Depression | Income | 16.2997 |
| Income | Life Events | 16.1404 |
| Depression | Stress | 14.5677 |
| BMI | Income | 13.9249 |
| BMI | Discrimination | 13.0348 |
| Depression | Medical Risk | 12.9667 |
| Discrimination | Income | 12.6559 |
| BMI | Life Events | 12.6086 |
| Income | Medical Risk | 11.8985 |
| Neighborhood Disadvantage | Life Events | 11.8888 |
| BMI | Depression | 11.8159 |
| BMI | Medical Risk | 11.6076 |
| Medical Risk | Life Events | 11.0932 |
| Neighborhood Disadvantage | Discrimination | 10.5255 |
| Discrimination | Medical Risk | 9.68267 |
| Age | BMI | 9.05879 |
| BMI | Education | 8.05817 |
| BMI | Stress | 7.62883 |
| Neighborhood Disadvantage | BMI | 7.14384 |
| Stress | Life Events | 7.09937 |
| Discrimination | Life Events | 6.85781 |
| BMI | Diet | 6.16898 |
| Income | Stress | 6.05402 |
| Age | Income | 5.90284 |
| Neighborhood Disadvantage | Income | 5.7389 |
| Discrimination | Education | 5.66232 |
| Age | Depression | 5.58917 |
| Neighborhood Disadvantage | Medical Risk | 5.12092 |
| Discrimination | Stress | 4.8111 |
| Neighborhood Disadvantage | Depression | 4.79894 |
| Diet | Income | 4.74904 |
| Education | Income | 4.45531 |
| Age | Life Events | 4.10359 |
| Diet | Life Events | 3.98751 |
| Neighborhood Disadvantage | Age | 3.60797 |
| Diet | Depression | 3.48525 |

| | | |
|---|---|---|
| Age | Medical Risk | 3.47825 |
| Neighborhood Disadvantage | Stress | 3.35339 |
| Neighborhood Disadvantage | Education | 3.15286 |
| Age | Discrimination | 3.10307 |
| Education | Depression | 3.09035 |
| Neighborhood Disadvantage | Diet | 2.85666 |
| Diet | Discrimination | 2.72128 |
| Education | Life Events | 2.70747 |
| Diet | Medical Risk | 2.33496 |
| Stress | Medical Risk | 2.2629 |
| Education | Medical Risk | 1.74381 |
| Age | Education | 1.69036 |
| Age | Diet | 1.49935 |
| Age | Stress | 1.45347 |
| Diet | Education | 0.881043 |
| Diet | Stress | 0.619795 |
| Education | Stress | 0.480053 |

**Table S1. All non-linear interactions.**
This is an expansion of the table included in Figure S2.
